# Use of face masks did not impact COVID-19 incidence among 10–12-year-olds in Finland

**DOI:** 10.1101/2022.04.04.22272833

**Authors:** Aapo Juutinen, Emmi Sarvikivi, Päivi Laukkanen-Nevala, Otto Helve

## Abstract

In fall 2021 in Finland, the recommendation to use face masks in schools for pupils ages 12 years and above was in place nationwide. Some cities recommended face masks for younger pupils as well. Our aim was to compare COVID-19 incidence among 10–12-year-olds between cities with different recommendations on the use of face masks in schools. COVID-19 case numbers were obtained from the National Infectious Disease Registry (NIDR) of the Finnish Institute for Health and Welfare, where clinical microbiology laboratories report all positive SARS-CoV-2 tests with unique identifiers in a timely manner, including information such as date of birth, gender, and place of residence. The NIDR is linked to the population data registry, enabling calculation of incidences. We compared the differences in trends of 14-day incidences between Helsinki and Turku among 10–12-year-olds, and for comparison, also among ages 7–9 and 30–49 by using joinpoint regression. According to our analysis, no additional effect seemed to be gained from this, based on comparisons between the cities and between the age groups of the unvaccinated children (10–12 years versus 7–9 years).

## Introduction

In fall 2021, the number of new COVID-19 cases was high globally [1]. In Finland, the delta variant had begun to spread in June, and by the end of July, delta was the dominant variant across the country. At that time, face mask use was recommended nationally in schools in children age 12 years and over. In some Finnish cities, this recommendation was extended to pupils age 10 years and above. The World Health Organization (WHO) stated that a risk-based approach should be applied to the decision to mask children between ages six and 11 years [2].

Our aim was to compare COVID-19 incidence among 10–12-year-olds between cities with different recommendations on the use of face masks in schools.

## Methods

COVID-19 case numbers were obtained from the National Infectious Disease Registry (NIDR) of the Finnish Institute for Health and Welfare, where clinical microbiology laboratories report all positive SARS-CoV-2 tests with unique identifiers in a timely manner, including information such as date of birth, gender, and place of residence [3]. The NIDR is linked to the population data registry, enabling calculation of incidences. Moving averages of 14-day incidences were used as a dependent variable in the statistical analysis. Estimated average percent changes (APC) were calculated in one-month periods. All figures were created using RStudio (R version 3.6.3) and all statistical analyses performed using the open source Joinpoint software (Joinpoint Regression Program, National Cancer Institute, USA, Version 4.9.0.0) as described previously [4].

Helsinki (population 661 887) and Turku (population 195 818) were selected for comparison, since the baseline incidence in the cities had been similar in August and September 2021. Helsinki implemented the national recommendation on face mask use at schools, while Turku had an extended recommendation that included those 10 years old and above.

## Results

We compared the differences in trends of 14-day incidences between Helsinki and Turku among 10–12-year-olds, and for comparison, also among ages 7–9 and 30–49, with the latter group representing the likely age group of the pupils’ parents. Moving averages of 14-day incidences and estimated average percentual changes (APC) are presented in Figure 1a. In August, there were no differences in APC values (difference, -0.1; *P*=.8). However, the APC was higher in September in Turku (difference, 2.9; *P*<.001), in October in Helsinki (difference, 2.3; *P*<.001), and in November in Turku (difference, -2.2; *P*<.001). The incidence for 7–9-year-olds was similar to that of 10–12-year-olds, but no such steep changes in November were observed in the incidence for 30–49-year-olds in either city (Figure 1b).

**Figure 1.**
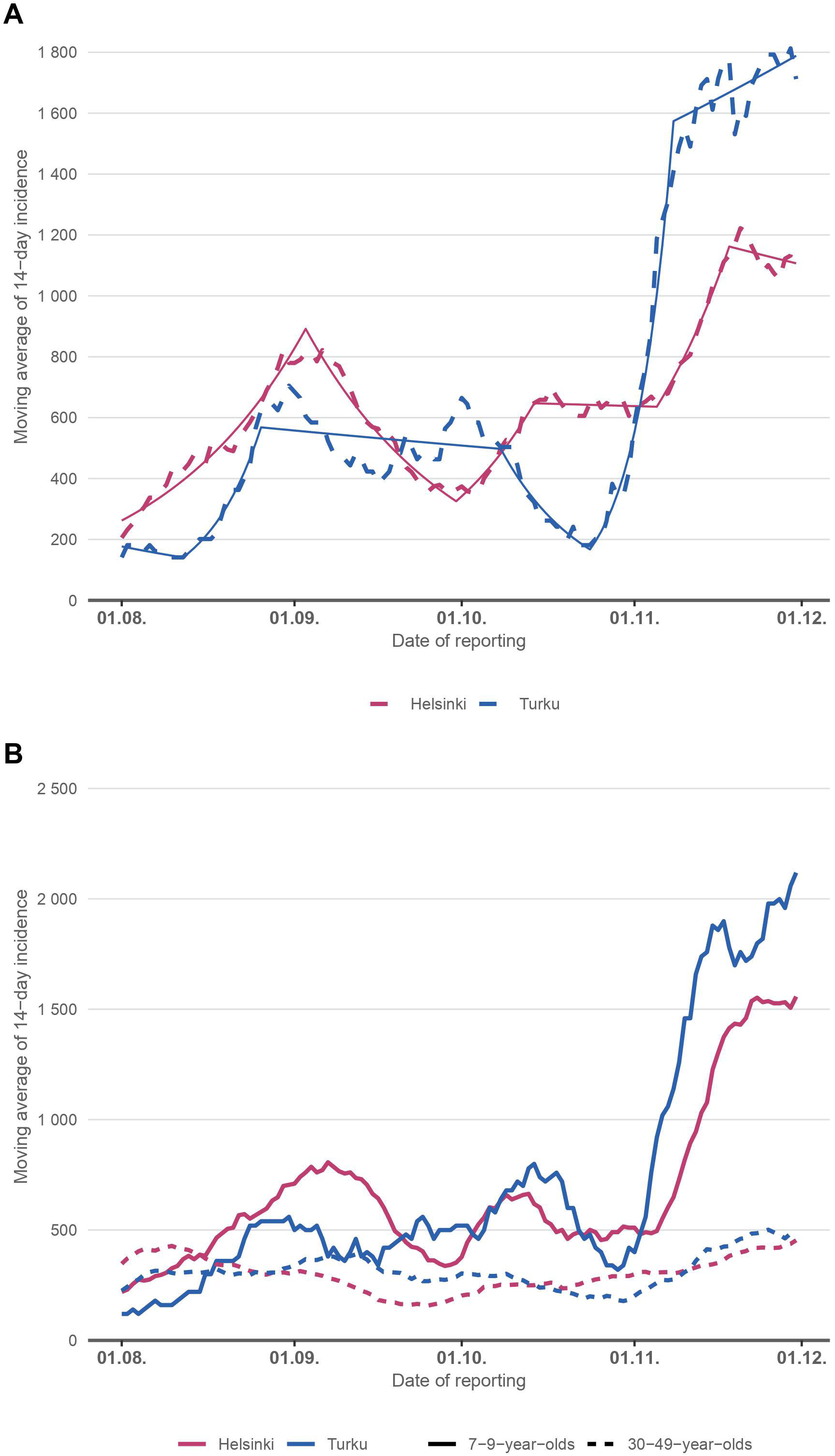
a) Moving average of COVID-19 incidence for 14 days (dashed line) and estimated APC values (solid line) in 10–12-year-olds in Helsinki (face masks not used in schools in this age group) and in Turku (face masks were used). b) Moving average of COVID-19 incidence for 14 days in 7–9-year-olds (solid line) and in 30–49-year-olds (dashed line) in Helsinki and Turku.

## Discussion

In fall 2021 in Finland, the recommendation to use face masks in schools for pupils ages 12 years and above was in place nationwide. Some cities recommended face masks for younger pupils as well, allowing us to assess the impact of face mask use in schools for younger pupils as a supplementary pandemic control measure. According to our analysis, no additional effect seemed to be gained from this, based on comparisons between the cities and between the age groups of the unvaccinated children (10–12 years versus 7–9 years).

The major limitation of our study is that schools are not the only place for children to have social contacts and be exposed to SARS-CoV-2. However, the lower incidence in vaccinated adults would indicate a lower risk of infection at home. Therefore, one would expect to see some differences in the age-specific incidences if masking was an effective way to control transmission in schools. Also, the timing for these observations was during a high circulation of the delta variant across the country. These results may not be valid during the omicron era.

## Data Availability

The data that support the findings of this study are available on request from the corresponding author on reasonable request.

## Acknowledgements

We are grateful to Claire Foley for proofreading the manuscript.

## Notes

### Competing Interest Statement

The authors have declared no competing interest.

### Funding Statement

This study did not receive any funding.

### Author Declarations

The Director of the Department for Health Security of Finnish Institute for Health and Welfare is the competent authority for assessing whether research requires institutional ethical review or if the Finnish communicable diseases law (Tartuntatautilaki 1227/2016) and the law on the duties of the Finnish institute for Health (Laki Terveyden ja hyvinvoinnin laitoksesta 668/2008) and Welfare allows the implementation of the research without seeking further ethical review. Based on his decision this research did not require further ethical review before implementation as its aim was to monitor the effectiveness of specific mitigation measures based on surveillance data (Tartuntatautilaki 1227/2016). The official document including the statement with signature, can be provided if necessary.

